# Excess Pneumonia Mortality During a Legionnaires’ Disease Outbreak in Flint, Michigan

**DOI:** 10.1101/19005942

**Authors:** ZO Binney, KN Nelson, AT Chamberlain

## Abstract

**Introduction:** From June 2014-October 2015, the Michigan Department of Health and Human Services (MDHHS) reported a Legionnaires’ disease (LD) outbreak in Genesee County, Michigan, of which Flint is the county seat. MDHHS’ final case count for the outbreak was 90 cases, including 10 deaths among Genesee County residents. As LD is not routinely tested for as a cause of community-acquired pneumonia, the size of the outbreak may have been underestimated. Specifically, some LD cases may have been classified as pneumonias of other or unexplained etiologies.

**Objective:** To estimate excess pneumonia deaths in Genesee County during the 2014-15 LD outbreak and compare this with reported deaths due to LD.

**Methods:** We used data from the CDC WONDER database, which contains monthly county-level counts of death certificates, on all pneumonia deaths among residents of Genesee and a set of similar counties from 2011-2017. We calculated excess pneumonia deaths in Genesee during the period of the LD outbreak (May 2014-October 2015). We used data from the Genesee County Vital Records Division to map pneumonia deaths by census tract to assess geographic overlap with reported LD cases.

**Results:** We estimated 70.0 excess pneumonia deaths (90% uncertainty interval (UI): 36 to 103) in Genesee County during the LD outbreak. This is substantially higher than the 10 LD deaths among Genesee County residents reported by MDHHS. Areas of high pneumonia mortality overlapped with those with high LD incidence and were primarily located in western Flint and northwestern non-Flint Genesee County.

**Conclusions:** These findings are consistent with the hypothesis that the LD outbreak was larger than reported. Earlier detection and response to this outbreak may have facilitated identification of these additional cases.

## Introduction

*Legionella pneumophila* is a common cause of community-acquired pneumonia and the leading cause of disease outbreaks due to drinking water in the United States.^1,2^ Infection with *Legionella* can lead to Legionnaires’ disease (LD), a severe form pneumonia.^3,4^ Many cases can be successfully treated with antibiotics, but LD is fatal in approximately 10% of cases.^4^

From June 2014-October 2015, the Michigan Department of Health and Human Services (MDHHS) reported a LD outbreak in Genesee County, Michigan, of which Flint is the county seat. Officially, the outbreak consisted of 90 confirmed LD cases, including 12 deaths (10 of whom were Genesee county residents).^5^ According to the National Outbreak Reporting System maintained by the Centers for Disease Prevention and Control and Prevention (CDC), the median size of LD outbreaks in the U.S. from 2009-2017 was 3 cases (IQR: 2–5) and 0 deaths (IQR: 0–1), making this the fourth-largest known LD outbreak since 2009.^6^

The timing of this outbreak coincided with the water crisis in the City of Flint that began in late April 2014, when treated water from Lake Huron supplied by the Detroit Water and Sewerage Department was replaced with locally-treated water from the Flint River.^7^ This change resulted in widespread effects on water quality primarily in Flint but also in greater Genesee County.^7,8^ One such effect of the switch was a reduction in the level of free chlorine present in the municipal water supply, a change which can be conducive to growth and persistence of *L. pneumophila*.^7,8^ Such alterations may have been linked to the LD outbreak, though its etiology remains unresolved.^7^

Despite a five-fold increase in LD incidence since 2000 and approximately 7,000 cases reported in 2017, LD is generally considered to be underdiagnosed.^5,9-11^ Indeed, a recent report from the National Academies of Sciences, Engineering, and Medicine (NASEM) estimated there may be 52,000-70,000 total LD cases per year.^11^

The CDC has recommended LD testing for specific groups, including individuals with pneumonia requiring ICU care or who failed initial antibiotic treatment, with recent travel histories, who are immunocompromised, or who are residents in a healthcare facility where a specific *Legionella* exposure risk has been identified.^4^ Even among these groups physicians may not routinely test for *Legionella* as a cause of pneumonia, in part because LD often responds to broad-spectrum antibiotics.^4^ For community-acquired pneumonias, physicians are generally encouraged to test for LD only when made aware of an outbreak.^9^ Indeed, the number of urine antigen tests for LD diagnosis conducted by Genesee County hospitals increased 45% from 2015 to 2016^12^ after the medical community was notified of the ongoing LD outbreak in Genesee County.

Because increased clinical testing for LD has often been contingent upon awareness of outbreaks, prompt communication from public health officials to local healthcare providers is critical for outbreak-associated case identification. Although the first official outbreak-associated LD cases in Genesee County were admitted to the hospital in June 2014, infection control personnel at local hospitals were not officially notified of an elevated risk of LD until February 2015, and the outbreak was not acknowledged publicly until January 2016.^13^ This timeline suggests the potential for outbreak-associated LD cases to have been misdiagnosed or missed entirely.

To determine whether additional LD cases may have gone unidentified during the 2014-15 Genesee outbreak, we used data on pneumonia deaths from 2011-2017 in Genesee and other comparable counties to 1) quantify whether and how many excess pneumonia deaths occurred in Genesee County during the outbreak, and 2) compare this number to the official LD death count reported. We also explored whether excess pneumonia mortality and LD cases occurred in similar geographic areas.

## Methods

### Data Sources

#### Pneumonia Deaths

We extracted pneumonia deaths from two sources: the CDC WONDER Multiple Cause of Death database and the Genesee County Vital Records Division (GCVRD).

From the CDC WONDER database we abstracted monthly counts of non-viral pneumonia and LD deaths (hereafter referred to as “pneumonia deaths”) in Genesee and control counties from 2011-2017.^14^ Because Genesee County is classified as a “Medium Metro” county according to the National Center for Health Statistics 2013 Urbanization Classifications, we selected all other Medium Metro counties (n = 45) from Michigan and four neighboring states (Illinois, Indiana, Ohio, and Wisconsin) as control counties.^15^ Pneumonia deaths were identified using ICD-10 codes J15-J18 for non-viral pneumonia and A48.1 and A48.2 for LD. In months with suppressed death counts for Genesee County (fewer than 10 deaths per month due to either pneumonia or LD) in WONDER, we assumed 8 such deaths during the month.

In addition to data from CDC WONDER, staff from the news program *Frontline* provided the authors with a dataset they compiled on all pneumonia (including LD) deaths in Genesee County from 2011-2017. This data was abstracted from death certificates obtained from GCVRD and included residential census tract information on all deaths.

We used the WONDER data to estimate excess pneumonia mortality and the GCVRD data to map pneumonia deaths since WONDER data does not include geolocation data beyond the county level.

#### LD Cases

*Frontline* compiled a list of LD outbreak cases by aggregating data from several sources: a list of 46 confirmed LD cases from Genesee County diagnosed between June 2014 and March 2015 provided by MDHHS, as well as 31 additional cases *Frontline* identified from legal documents and news reports. This list included 67 total cases. *Frontline* provided the authors an anonymized version of this case list with the residential census tract of each case. From this list, we excluded 9 cases who were non-Genesee residents and 3 with an unknown hospitalization date. The remaining 55 cases were used in the mapping analysis.

Of note, MDHHS’ final report listed 90 confirmed cases and 12 deaths, suggesting *Frontline*’s list was missing 23 cases.^2^ The MDHHS report only included the cases’ months of symptom onset, however, and did not contain the geographic information necessary for case mapping.^2^ Of the 12 deaths in the MDHHS report, 10 were Genesee County residents.

#### Population Data

To calculate incidence and mortality rate denominators we gathered county- and census tract-level population data from the Census and American Fact Finder for 2010-2017. We assumed a simple linear population change between mid-year (July 1) estimates for all counties.

### Statistical Analysis

#### Excess Deaths Estimation

We used a Bayesian negative binomial model to model monthly counts of pneumonia deaths in Genesee and control counties with an offset of the natural log of the county population in a given month. Our data consisted of 84 monthly counts of pneumonia deaths over 7 years in each of two geographical areas: Genesee County, and the control counties described above (n = 168 total observations). The model included fixed effects for Genesee vs. control counties, month, and flu season year (defined as October-September) to control for secular trends in pneumonia incidence and influenza season severity. Because Genesee pneumonia mortality was particularly elevated during May-October 2014, we included a term indicating the outbreak stage: “early” outbreak (May 2014-October 2014), “late” outbreak (November 2014-October 2015), or not during the outbreak (all other months from 2011-2017). Although the first LD case was not reported until early June 2014, we chose May 2014 as the start of the outbreak in our main analysis to capture any deaths that may have occurred earlier. We also included an interaction between the location and outbreak stage terms. The interaction terms for *early outbreak x Genesee* and *late outbreak x Genesee* represent the estimated difference in the differences in pneumonia mortality between Genesee and control counties during versus outside the LD outbreak. In other words, they represent an estimate of the change in pneumonia mortality in Genesee during the outbreak as compared to non-outbreak periods.

We used weakly regularizing priors for the intercept [N(−10, 5)], fixed effect coefficients [N(0, 1)], and the shape parameter [Gamma(0.01, 0.01)]. The model’s linear predictor represents the natural log of the number of pneumonia deaths for each month and location. We calculated predicted pneumonia mortality rates by exponentiating this value and dividing by the population for that location and month to yield a predicted rate.

To estimate excess deaths we ran a second model, excluding the outbreak and outbreak x Genesee terms, on the 150 observations unaffected by the LD outbreak (i.e. all control county months as well as Genesee months outside the LD outbreak). We sampled from this second model’s posterior predictive distribution to generate 4,000 counterfactual predictions for the expected number of deaths in Genesee each month from May 2014-October 2015 absent the LD outbreak. For each posterior prediction we took the difference between the prediction and the observed death count for that month, summed these differences across the 18 months of the outbreak (May 2014-October 2015), and used the resulting distribution of summed differences to calculate a mean and 90% uncertainty interval (UI) for excess deaths.

#### Mapping

To map the residential census tracts of LD cases and pneumonia deaths, we obtained county and census tract shapefiles (2010 Census version) from the U.S. Census Bureau. We calculated and mapped the LD incidence during the “early” outbreak period (June 2014 – October 2014), and the entire duration of outbreak (June 2014 – October 2015) by census tract in Genesee County. We mapped pneumonia mortality by residential census tract across those same time periods and two additional periods: before the outbreak (January 2011 – May 2014) and after the outbreak (October 2015 – December 2017). We compared areas of high LD incidence with those of high pneumonia mortality during the outbreak, and also identified census tracts with the largest changes in pneumonia mortality before as compared to during the LD outbreak.

#### Sensitivity Analyses

We conducted sensitivity analyses to assess the robustness of our results to several key assumptions. First, we calculated excess deaths assuming that the outbreak began in June (to mirror official reports), rather than May. Second, we considered an excess deaths model with a fixed effect for calendar year 2017, because our data showed that pneumonia deaths were elevated in Genesee relative to control counties during this period for reasons that are unknown but likely unrelated to the 2014-15 LD outbreak. Lastly, to allay concerns about small counts of LD cases and pneumonia deaths by census tract, we also mapped LD and pneumonia mortality by pseudo-quadrants in Flint and surrounding Genesee County rather than by census tract.

#### Data and Code Availability

All data, code, and output for our models, maps, and sensitivity analyses is <u>available on Github</u>.

All analyses were done in R v.3.5.1; the models were fit using the brms package and mapping was performed using the sp package.

This study was reviewed and approved by the Institutional Review Board at Emory University.

## Results

Using the CDC WONDER database, we identified 1,461 pneumonia deaths among Genesee County residents and 24,692 pneumonia deaths among residents across 45 control counties from 2011-2017. Deaths specifically coded with LD as a cause of death represented less than 0.2% of total pneumonia deaths. A complete list of control counties located in Michigan and surrounding states of Indiana, Illinois, Ohio, and Wisconsin is available (Supplemental Table 1).

The GCVRD data included 1,761 pneumonia deaths among Genesee County residents from 2011-2017, 1,751 of whom had residential census tract information. While the GCVRD data contained 348 more pneumonia deaths than were identified via WONDER, the GCVRD and WONDER data showed similar seasonal and secular trends.

### Pneumonia Mortality Visualization

Monthly pneumonia mortality rates from 2011-2017 showed an increase in pneumonia mortality beginning in May 2014 in Genesee as compared to control counties, coincident with the start of outbreak-associated LD cases (Figure 1).

**Figure 1.**
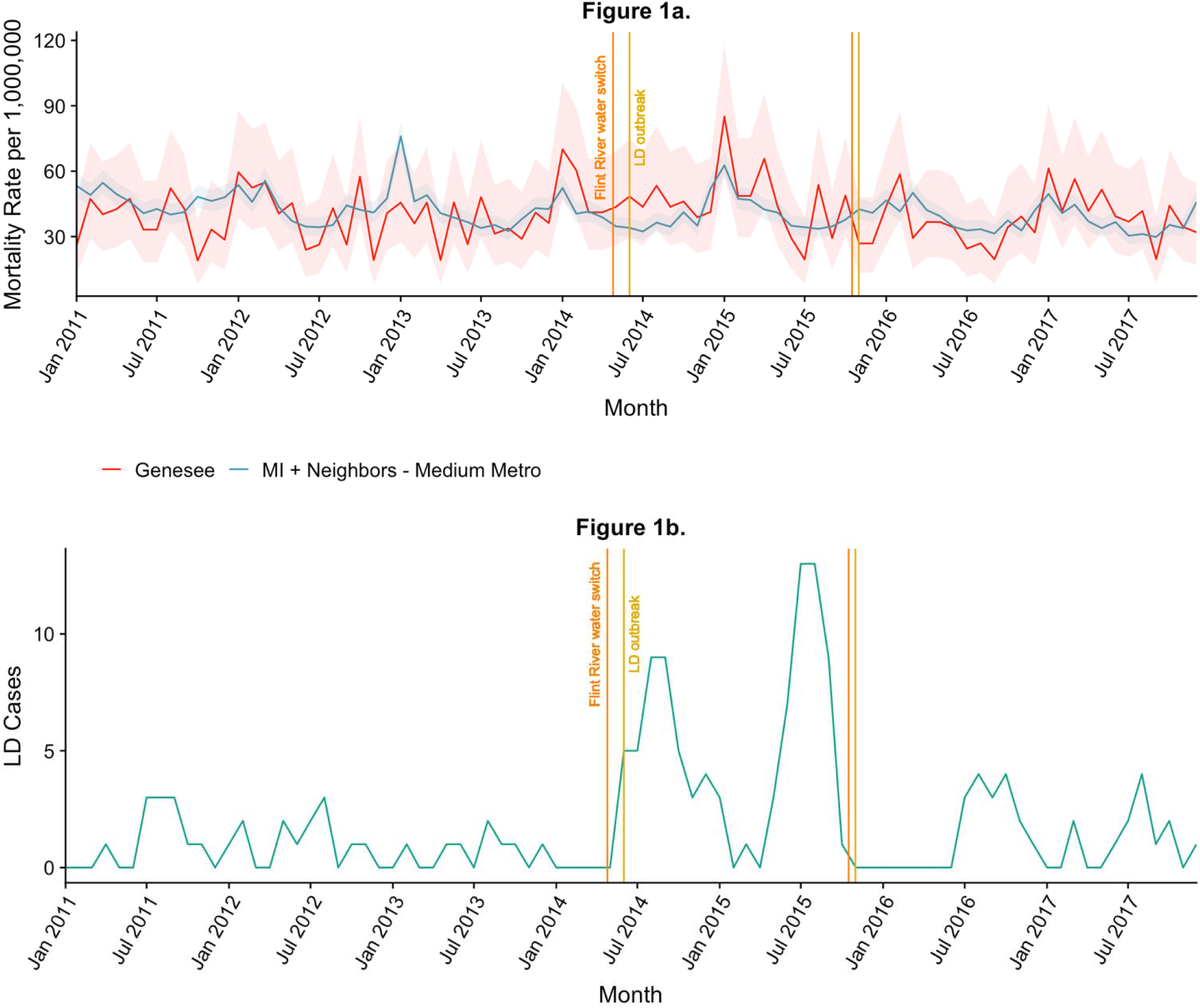
Pneumonia Mortality and LD Outbreak Cases, 2011-2017, Genesee and control counties. **A**. Pneumonia mortality in Genesee and control counties with exact 95% Poisson confidence intervals. The vertical orange lines delineate the start and end of the Flint municipal water source change from Detroit to the Flint River (April 25 2014 – October 16 2015); the vertical yellow lines delineate the official start and end of the Flint LD outbreak per MDHHS (June 1 2014 – October 31 2015). **B**. The outbreak curve of LD cases, by month of symptom onset. This includes the 90 confirmed LD cases reported by MDHHS.

### Excess Deaths Estimation

Fit and convergence diagnostics, including R-hat measures, trace plots, and posterior predictive checks suggested the Bayesian negative binomial models converged and fit well (see the <u>GitHub page</u>).

The pneumonia mortality rate was 6% lower in Genesee County than control counties during months before and after the LD outbreak (rate ratio (RR) 0.94, 90% UI: 0.88-0.99). During the LD outbreak, Genesee County pneumonia mortality was 31% (RR 1.31, 90% UI: 1.10-1.54) and 13% (RR 1.13, 90% UI: 0.96-1.32) higher in the outbreak’s early and late periods, respectively, as compared to control counties (Table 1).

**Table 1.**
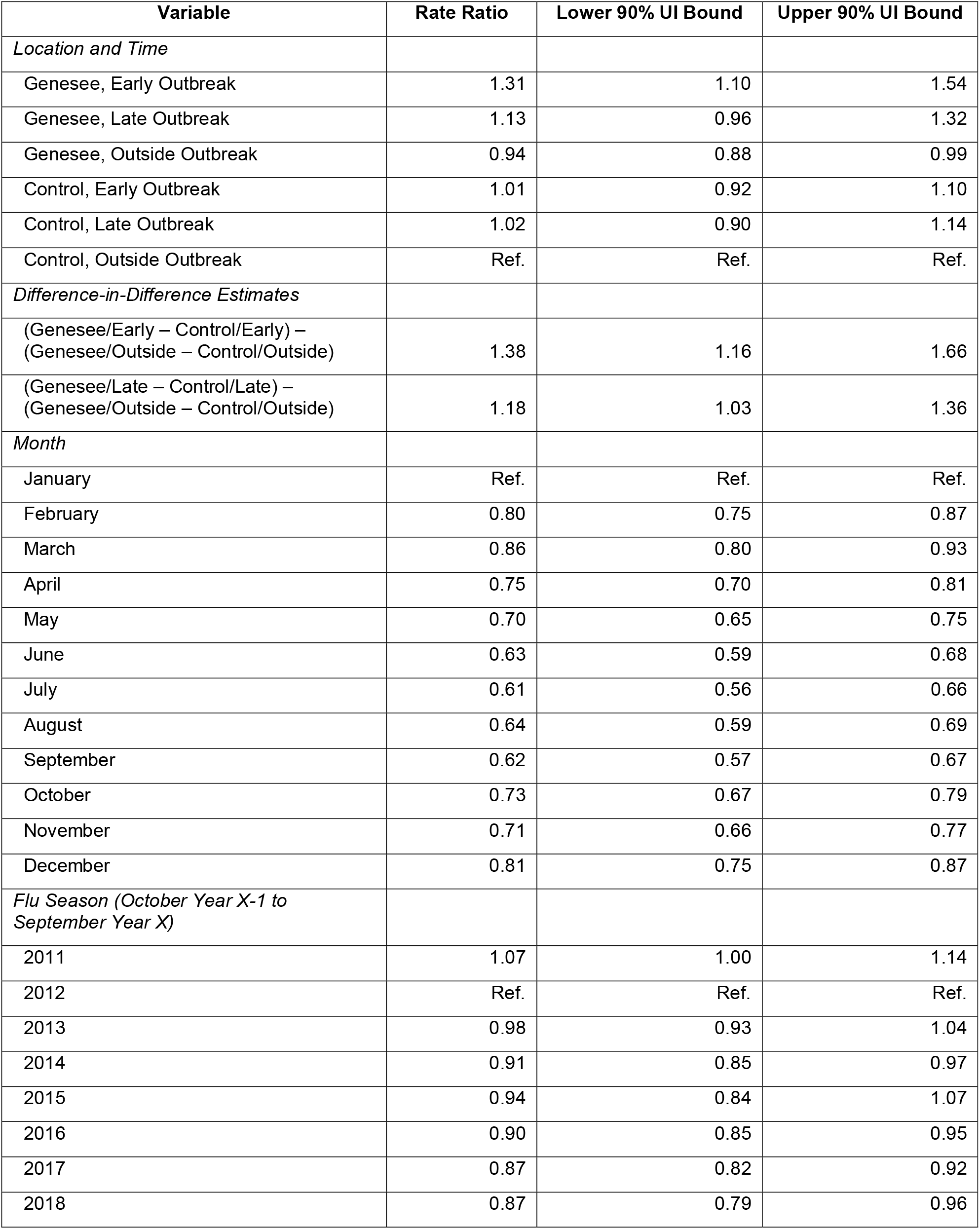
Model-based pneumonia mortality rate ratios for Genesee County compared to control counties with 90% uncertainty intervals.

Controlling for monthly and yearly incidence trends, pneumonia mortality increased 38% (RR 1.38, 90% UI: 1.16-1.66) and 18% (RR 1.18, 90% UI: 1.03-1.36) in Genesee County during the LD outbreak relative to the periods before and after the outbreak.

Of note, the estimated pneumonia mortality rate is lower in Genesee County prior to and after the outbreak but substantially elevated during the outbreak, particularly during the early outbreak period from May–October 2014. (Figure 2).

**Figure 2.**
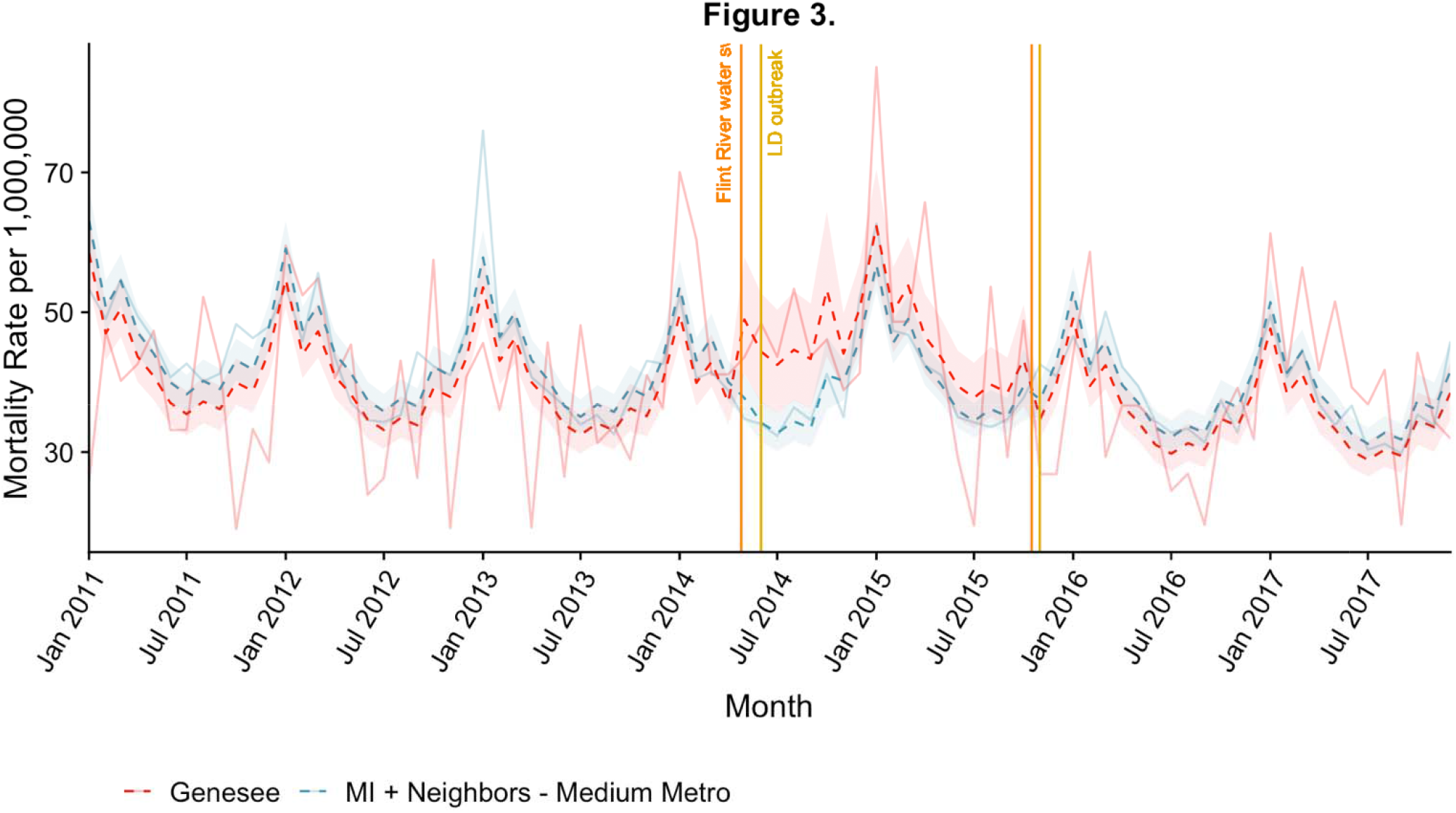
Plot of fitted pneumonia mortality rates for Genesee and control counties, 2011-2017. Monthly pneumonia mortality rates (solid lines) alongside the model’s predictions (dotted lines) for Genesee (red) and control counties (blue). 90% uncertainty intervals for model-estimated expected mortality rates for Genesee and control counties are shown in the corresponding color.

Finally, we estimated 70.0 (90% UI: 36-103) excess pneumonia deaths among Genesee County residents during the LD outbreak. (Figure 3)

**Figure 3.**
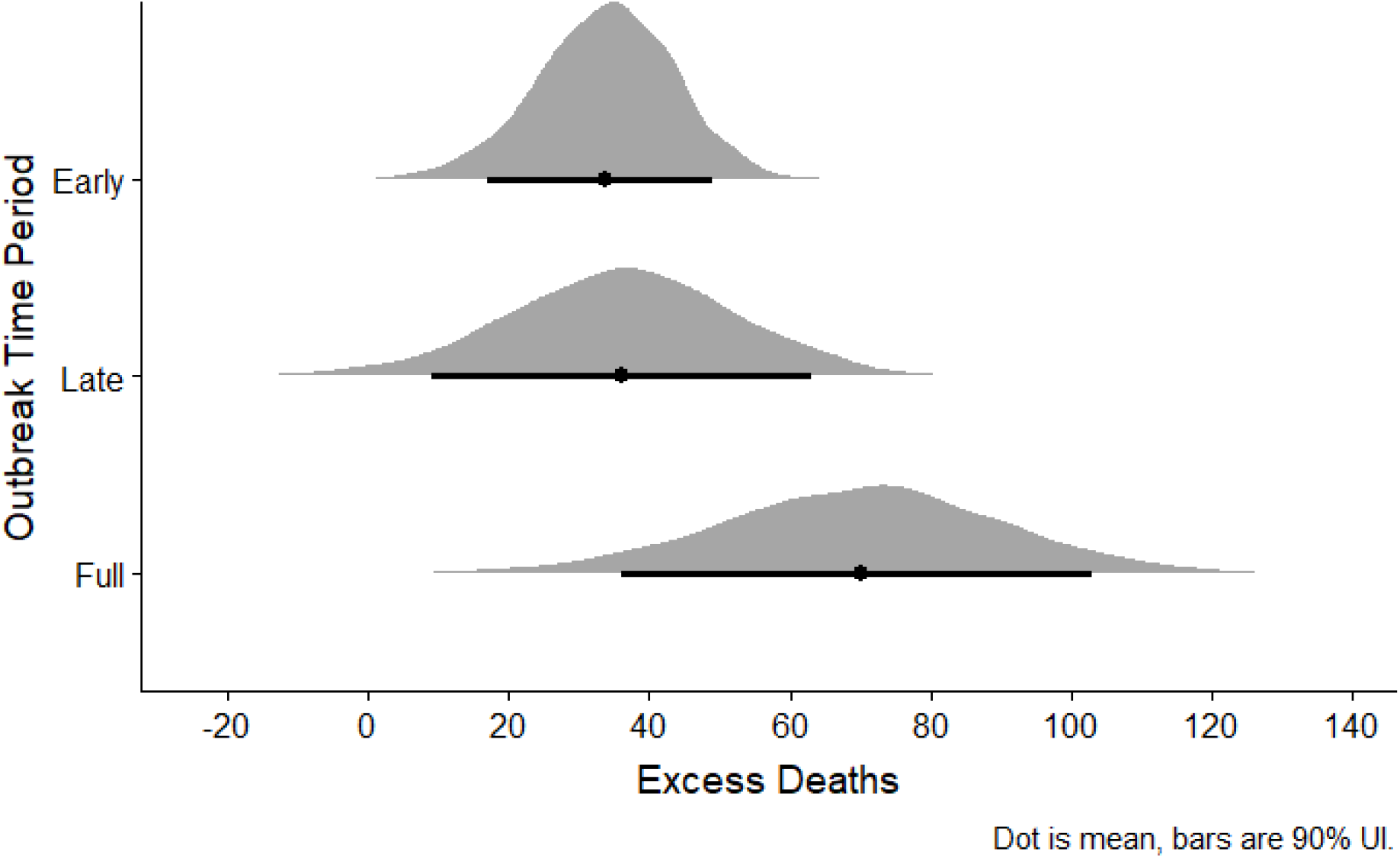
Posterior predictive distribution of excess deaths in the early (May-October 2014), late (November 2014-October 2015), and full (May 2014-October 2015) outbreak periods.

### Mapping

Areas with high LD incidence were concentrated in western Flint and in northwestern, non-Flint Genesee County. (Figure 4) These same areas had the highest increases in pneumonia mortality during the time period of the LD outbreak relative to before the outbreak. (Figure 5)

**Figure 4.**
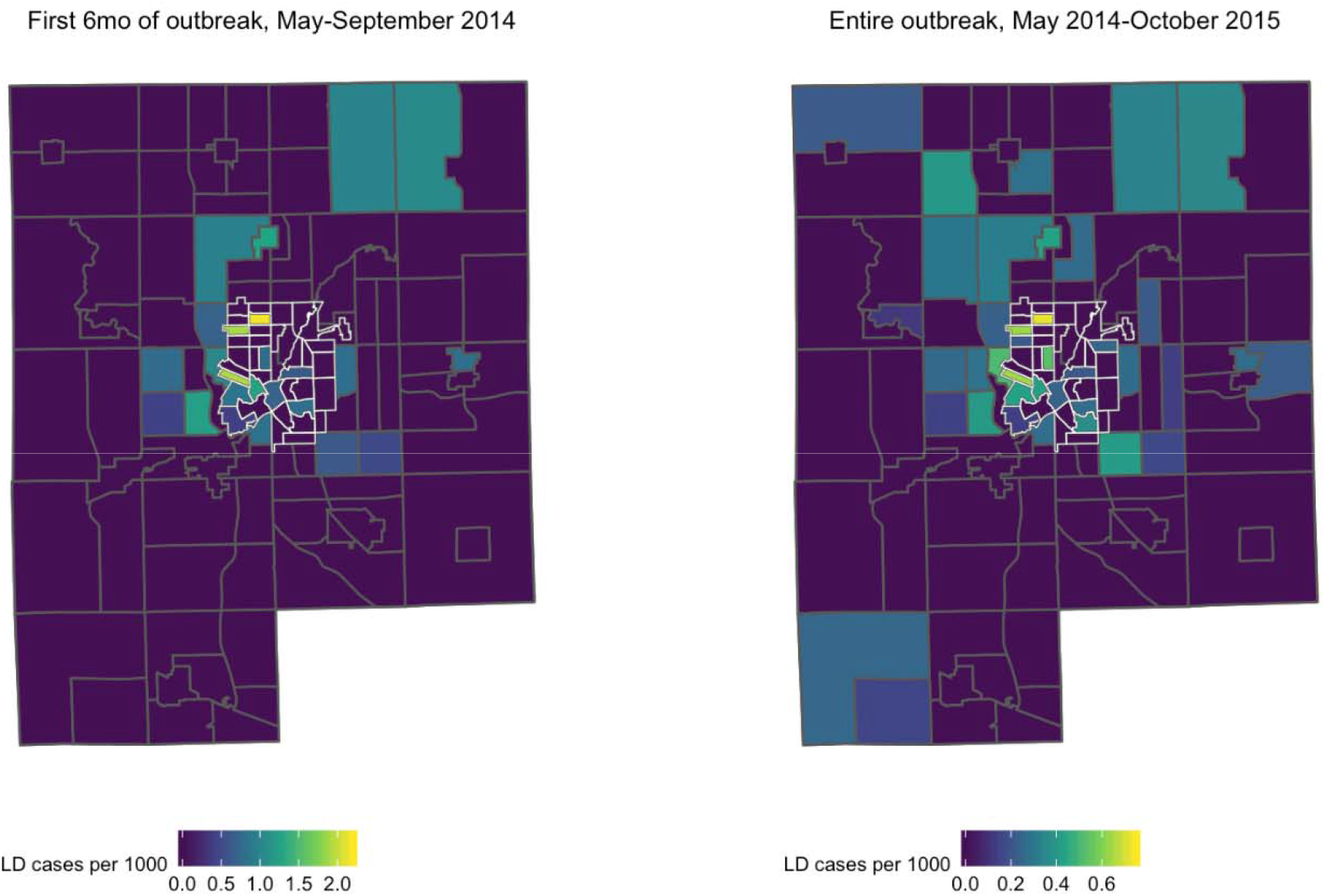
Legionnaires’ disease incidence by census tract in Genesee County, Michigan, 2011-2017. Census tracts with grey borders are non-Flint Genesee County; census tracts with white borders are located in the city of Flint. Each tract is colored based on LD incidence (cases per 1000) during the specified time period. A. Census tracts in green and yellow indicate high rates of LD during the first 6 months of the outbreak (left, May 2014 – October 2014) and over the course of the entire outbreak (right, May 2014 – October 2015). Scales are different to emphasize census tracts with high incidence during each time period.

**Figure 5.**
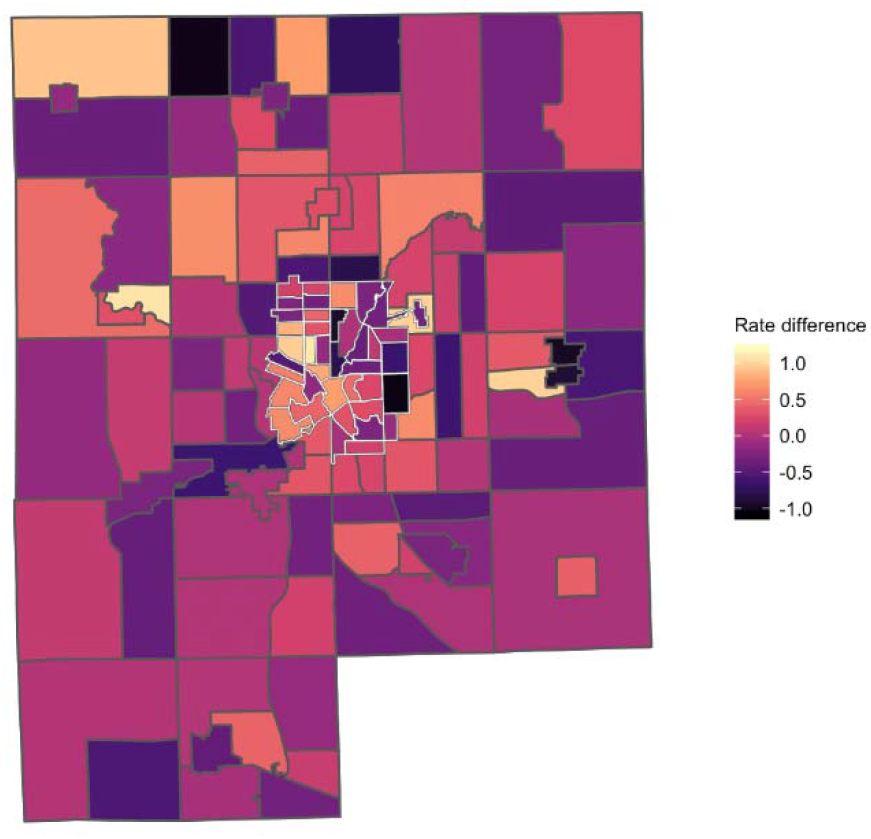
Difference in pneumonia mortality during as compared to before the LD outbreak in Genesee County, Michigan, 2011-2015. Census tracts with grey borders are non-Flint Genesee County; census tracts with white borders are located in the city of Flint. Each tract is colored based on the change in pneumonia mortality rate (cases per 1000) during as compared to before (January 2011 – April 2014) the outbreak. Positive values indicate higher mortality during the LD outbreak than before the outbreak; census tracts with the most positive values are shown in increasingly lighter shades of orange and yellow.

### Sensitivity Analyses

None of the sensitivity analyses we conducted substantively affected our conclusions. Considering June, rather than May, as the beginning of the outbreak reduced our excess deaths estimate to 64.7 (90% UI: 32-96) (<u>see Github</u>). Including a separate fixed effect for calendar year 2017 and an interaction with Genesee increased our estimate of the excess pneumonia deaths from 70.0 to 80.4 (90% UI: 47-112) (<u>see Github</u>). This model also provides a better fit, particularly for pneumonia mortality in 2017 in Genesee. Assuming 5 deaths instead of 8 for months with suppressed counts in WONDER did not change the estimate substantively. Lastly, pseudo-quadrant maps did not show meaningfully different trends from those showing census tract-level data (<u>see Github</u>).

## Discussion

We estimated 70.0 excess pneumonia deaths (90% UI: 36-103) in Genesee County during the LD outbreak. This is substantially higher than the 10 LD deaths among Genesee County residents reported by local public health officials. Areas with high pneumonia mortality overlapped those with high LD mortality, particularly in western Flint and northwestern non-Flint Genesee County. Taken together, this evidence suggests that the excess deaths we estimated may be partially attributable to unidentified LD.

Our results are consistent with other recent LD analyses attempting to estimate the true burden of the disease. A recent analysis from Connecticut estimates that only 10.6% of hospitalized LD cases are diagnosed – suggesting the total hospitalized cases could be 9.4 times reported totals.^16^ The 2019 NASEM report estimates the total LD burden to be upwards of 50,000 cases annually, implying that the actual number of LD cases in the U.S. could be 7-10 times higher than official reports.^11^

Identifying every case in an outbreak is often impossible given the practical limitations of outbreak investigation and the challenges of retrospective case identification. As such, it is not surprising that some outbreak-associated LD cases would go undetected. However, prompt communication with local communities and healthcare professionals – such as via a Health Alert Network (HAN) or another platform for rapid and broad distribution of public health information – is a key component of an LD outbreak response.^17^ This communication raises awareness among physicians of the potential for *Legionella* exposure and may encourage them to test for LD as a cause of pneumonia. Notifying the community of an LD outbreak also alerts the public to their own risk of exposure, allowing additional cases to seek care and be diagnosed. Compared to other recent outbreaks of community-acquired LD, information regarding this outbreak was communicated to stakeholders relatively late: although the first cases were diagnosed in June 2014, local infection control personnel were not officially notified for another 8 months (February 2015), and the public was not notified for another 19 months (January 2016), after the outbreak had ended.^13^ This timeline stands in stark contrast to, for example, a large LD outbreak that occurred in the South Bronx in New York City in 2015.^17^ In this outbreak, clinicians at local hospitals were notified by the health department within a week of the detection of a cluster of LD cases. Physicians were encouraged to test for *Legionella* in patients with suggestive respiratory symptoms and request that autopsies be performed on patients with unexplained respiratory illness. The general public was notified within weeks of the first cases and community engagement activities were coordinated to allay community concerns about the outbreak and to identify additional cases. As a result of these efforts, the South Bronx outbreak was declared over in August 2015 – less than two months after it had started.

It is important to note that publicizing information regarding a cluster of LD cases, especially in its early stages, is a sensitive decision that carries risks as well as benefits. No two outbreaks are exactly the same, and public health officials must rely on their experience and their assessment of the specific contours of the situation in order to decide when and how much information to release to the public. Moreover, in this particular case, local public health officials were simultaneously devoting resources to addressing other health effects of the water crisis. Even so, it remains difficult to reconcile the severe delays in communication that occurred in response to the 2014-15 LD outbreak in Genesee County. It is possible that these delays prevented identification of additional cases and deaths that may have facilitated earlier resolution of the outbreak.

Our analysis has several limitations. First, it is unlikely that all 70 excess pneumonia deaths we identified were due to LD. Some deaths may have been due to other causes of pneumonia, though we excluded viral pneumonias to mitigate this risk. Perhaps less likely, some individuals who died of LD may not have had pneumonia listed as a cause on their death certificate; these cases would not have been included in our analysis.

Second, our analysis is limited to estimating outbreak-associated LD deaths; it did not allow us to draw direct conclusions about the total number of non-lethal LD cases. That said, it is reasonable to assume the fraction of unidentified LD cases may be similar to or larger than that for unidentified deaths, assuming that fatal cases were more likely to have been identified.

Third, our model rests on the assumption that the control counties we selected are similar to Genesee with respect to any factors that might influence risk of pneumonia and LD – that is, they represent what would have happened in Genesee absent the LD outbreak. In addition to choosing counties that had a similar urbanicity to Genesee, we chose counties from the surrounding region to control for climate-related factors associated with pneumonia acquisition, including the impact of geography on influenza seasonality and spread. Of note, differences in age, race, and other sociodemographic features between Genesee and control counties should not bias our results unless these differences changed over the course of our study (2011-2017); we consider this implausible. Furthermore, the trends in mortality in the two groups before the outbreak, while unstable due to relatively small numbers of pneumonia deaths in Genesee, were not obviously different.

Finally, we used the Multiple Cause of Death database from WONDER to identify pneumonia deaths in Genesee County, which includes the underlying cause of death in addition to any other causes of death listed on the death certificate by the physician. Although this may have captured some individuals for whom pneumonia, and unidentified LD, was not their *primary* cause of death, we chose this approach to ensure an inclusive case definition. Moreover, because mortality from LD is greater among individuals with pre-existing health conditions than among healthy persons^1^, it is especially likely that LD cases would have had a non-pneumonia underlying cause of death listed.

Prompt and thorough response to outbreaks of LD is important beyond Genesee County. LD incidence has increased dramatically in the U.S. since 2001, which may reflect the changing epidemiology of LD.^1^ Both the increased prevalence of LD risk factors (old age, immunosuppression) and a higher proportion of the population living in cities with aging and centralized water systems may be among key factors increasing LD risk. As these risk factors become increasingly common, implementing routine testing of patients hospitalized with pneumonia for LD can help to rapidly identify and investigate LD outbreaks as well as provide critical data for further understanding the epidemiology of this disease.^11^ This information can ultimately inform regulatory policies aimed at protecting the public against *Legionella* in water systems, such as those implemented by New York City in response to the South Bronx outbreak.^11,18^ These policies should be informed by a strong evidence base of research into both individual-level risk factors for severe disease as well as the environmental and infrastructural conditions that put populations at high risk.

In summary, we found approximately 70 excess pneumonia deaths during the 2014-2015 outbreak of LD in Genesee County, Michigan. These deaths were located in similar geographic areas to reported LD cases. Given that there were only 10 reported LD deaths among Genesee County residents, our findings are consistent with the hypothesis that there were additional unidentified LD deaths and cases during this time period. Though all reported cases were investigated by public health officials, delays in communication may have resulted in a failure to identify others. Prompt and complete identification of cases can inform epidemiologic investigations aiming to identify common exposures as well as provide critical information about environmental and structural risk factors for LD.

## Data Availability

All data, code, and output for our models, maps, and sensitivity analyses is available on Github.

https://github.com/zbinney/Flint_Legionnaires

## Acknowledgements

The authors wish to thank Dr. Ruth Berkelman for her advice and feedback; Kayla Ruble and Jacob Carah from *Frontline* for providing case mapping data; and Drs. Emily Ricotta (Epidemiology Unit, Laboratory of Clinical Immunology and Microbiology, NIAID, NIH) and Jon Fintzi (Biostatistics Research Branch, NIAID, NIH) for their generous feedback and statistical review.

## Supplemental Material

**Supplemental Table 1.** List of selected control counties in MI, IL, IN, OH, WI

| County | State |
| --- | --- |
| Cass | Michigan |
| Clinton | Michigan |
| Ingham | Michigan |
| Eaton | Michigan |
| Kalamazoo | Michigan |
| Van Buren | Michigan |
| Washtenaw | Michigan |
| Boone | Illinois |
| Henry | Illinois |
| Marshall | Illinois |
| Mercer | Illinois |
| Peoria | Illinois |
| Rock Island | Illinois |
| Stark | Illinois |
| Tazewell | Illinois |
| Winnebago | Illinois |
| Woodford | Illinois |
| Allen | Indiana |
| Posey | Indiana |
| St. Joseph | Indiana |
| Vanderburgh | Indiana |
| Warrick | Indiana |
| Wells | Indiana |
| Whitley | Indiana |
| Carroll | Ohio |
| Fulton | Ohio |
| Greene | Ohio |
| Lawrence | Ohio |
| Lucas | Ohio |
| Mahoning | Ohio |
| Miami | Ohio |
| Montgomery | Ohio |
| Portage | Ohio |
| Stark | Ohio |
| Trumbull | Ohio |
| Wood | Ohio |
| Brown | Wisconsin |
| Columbia | Wisconsin |
| Dane | Wisconsin |
| Douglas | Wisconsin |
| Green | Wisconsin |
| Iowa | Wisconsin |
| Kewaunee | Wisconsin |
| Oconto | Wisconsin |

